# Pre-existing health conditions and severe COVID-19 infection: Analysis of commercial health insurance data from 690,000 infected patients

**DOI:** 10.1101/2021.03.11.21252708

**Authors:** Nathan E. Wineinger, Victoria Li, Jill Waalen, Eric J. Topol

## Abstract

The development and distribution of new vaccines promises an end to the COVID-19 pandemic. With the elderly and front line workers first in line, the vaccination strategy for the general population remains unclear. In this study we identified 690,000 patients infected with COVID-19 with commercial health insurance coverage across the country between April 1, 2020 and September 30, 2020. From prior health care claims, we determined each person’s pre-existing diseases among 26 common chronic diseases. Across age-sex strata we determined the relationships between these conditions and severe COVID-19 infections: ICU admission and extended in-patient stay. We classify disease according to risk, develop multivariable models to predict infection outcomes, and created an online risk assessment tool to estimate risk of ICU admission based on pre-existing health conditions. Our results could be used to help guide risk, vaccination health policy, and personal decision-making as these become available to the general population.

## Introduction

Coronavirus disease 2019 (COVID-19) was first detected in the United States in January 2020. Since then, nearly 26.2 million confirmed cases and 512,000 deaths due to COVID-19 have been reported in the United States (as of March 1, 2021).^1^ Early data from cohort studies and case series have identified potential risk factors for developing severe disease, including the association of comorbidities with hospital outcomes such as intensive care, mechanical ventilation, or in-hospital death.^2–4^ According to the Centers for Disease Control and Prevention (CDC), these risk factors include older age^5,6^ and other comorbidities, such as diabetes, chronic obstructive pulmonary disease, chronic kidney disease, hypertension, and immunocompromised states.^7,8^ Previous studies have developed risk scores to predict ICU admission and mortality using presenting clinical characteristics, but are limited in size or geographically.^9–11^

The rapid development of COVID-19 vaccines^12–14^ with very high efficacy and safety is poised to go down as one of, if not the greatest achievement in modern medicine. Subsequently, the phased allocation of vaccine distributions are designed to protect the most vulnerable in the population first (i.e., elderly and those with significant risk) and the health workers and first responders that administer care.^15^ As of March 1, 76.9 million vaccine doses have been administered in the U.S., with 1-2 million doses per day.^1^ Vaccine distribution is rapidly approaching Phase 2 – individuals in society with a high risk of exposure – and will hopefully begin being available to the general population in the coming months. With a population of about 331 million, even with the introduction of more vaccines and an increase in vaccine production, many will be waiting their turn (and some may elect to forego inoculations). While it is understood that the risk of severe infection is much lower in younger individuals and those without pre-existing conditions, little guidance is available for individuals at mild to moderate risk. Furthermore, there is a range of efficacy in the results from the six vaccine Phase 3 trials and a limited short-term vaccine supply, prompting many to strategize who should get the more potent vaccines, who might benefit from a single, primer dose with delay for a second, and how to optimize both the vaccines available and public health outcomes.

In this study, we set out to identify the clinical characteristics associated with severe COVID-19 infection among 690,000 individuals infected with COVID-19 between April 1, 2020 and September 30, 2020 with commercial health insurance from Blue Cross Blue Shield companies across the country. From health insurance claims data, we identified two primary outcomes: ICU admission related to COVID-19, and extended hospital stay due to the infection. The primary objective of this study was to develop risk models for predicting these outcomes from a geographically diverse group of patients hospitalized for COVID-19, representative of the general population that would be targeted for vaccine administration in the later phases of the distribution programs. We created an online COVID-19 risk assessment calculator based on these models that can be used to estimate risk based on pre-existing health conditions. Our results can provide insight into public health policy and potential for personal decision-making related to vaccinations.

## Methods

BCBS Axis, a data resource for health care claims, provider, and cost data that represents 36 Blue Cross Blue Shield and independent companies across the country was used in this analysis. The database includes de-identified health insurance claims data for over 87 million concurrent Blue Cross Blue Shield members across the United States and territories from the prior 40 months, in addition to a multitude of other insurance-related data. This database is accessed through a secured remote desktop application using two-factor authentication, and this system was created and is maintained by BCBSA.

We first gained access to this data in June 2020, and the database was updated monthly to reflect new claims. There is approximately a 2 month lag between updates to this database and the data available in claims. At the time of preparation of this manuscript, the database was last updated in late December 2020 and includes claims through October 2020. There is an additional lag of incomplete data within the claims of approximately 1 month (i.e., additional October claims will be included in the January 2021 update). And so, we elected to only include claims through September 2020 for all analyses.

Beginning on April 1, 2020, the Center for Disease Control and Prevention issued new ICD-10-CM coding and reporting guidelines for COVID-19.^16^ Confirmed positive diagnoses and presumptive positive tests were to be assigned ICD-10 code U07.1 (both coded as confirmed). Prior to this date, an imprecise combination of codes related to pneumonia, acute bronchitis, lower respiratory infection, acute respiratory distress syndrome, and other signs and symptoms related to COVID-19 (e.g., fever) were used. While some claims prior to this date in the BCBS Axis were recoded to reflect these new reporting guidelines, this was incomplete. And so, here we present only cases of confirmed COVID-19 via U07.1 ICD-10-CM Principal Diagnosis Code beginning on April 1 through September 2020.

Masked patient identifiers from all cases of confirmed COVID-19 discovered in all health claims – in-hospital (including inpatient and emergency room) and outpatient (including office visits and urgent care) – during this time period were extracted from the NDW database. ICU admissions were recorded using revenue codes 0200, 0201, 0202, 0203, 0204, 0206, 0207, 0208, and 0209. For admitted patients, the maximum length of stay was calculated from the first and last service dates. For all COVID-19 confirmed patients, a health history was generated based on the 26 common chronic diseases defined from the Chronic Conditions Data Warehouse^17^ (Alzheimer’s disease was merged with Alzheimer’s disease, related disorders, and senile dementia) as noted in ICD-10-CM Principal Diagnosis Codes from all claims data. Birth date (to determine age at diagnosis) and sex were extracted from a member information table. No data on race/ethnicity or socioeconomic status were available on individual data. No coverage length limits were used to exclude patients.

Patients were stratified by sex and age at first claim with confirmed COVID-19 infection: 18-35, 35-45, 45-55, 55-60, 60-65, 65-70. Patients above 70 were omitted. Two Boolean primary outcomes were considered: 1) ICU admission occurring on or after the first date of confirmed COVID-19 infection; and 2) a hospital length of stay greater than 1 day. Death cannot be inferred directly from these claims. In strata where the number of patients with one of these outcomes and chronic disease was not at least 20, this disease was excluded from analyses. Marginal associations between each of these outcomes and health history of each chronic disease were performed using logistic regression in each stratum. Multivariable analysis on each outcome using each chronic disease that met criteria as predictors was likewise performed using multivariable logistic regression in each stratum. Additionally, pairwise interactions between chronic kidney disease, diabetes, hyperlipidemia, and hypertension (the four most common conditions) were included in the multivariable models.

## Results

The Blue Cross Blue Shield Association (BCBSA) manages health insurance claims and related data from 36 Blue Cross Blue Shield and independent companies across the United States and territories within BCBS Axis. This represents over 86 million concurrent members with commercial health care insurance. Nearly 4 million claims with an ICD-10-CM Principal Diagnosis Code for confirmed COVID-19 (U07.1) were identified from all claims in the NDW. This includes 690,173 unique patients that met study inclusion criteria with a health care claim for COVID-19 between April 1, 2020 and September 30, 2020. Age and sex breakdown of this study cohort is included in Table 1.

**Table 1.** Total sample size (N) for each age-sex strata, and number of individuals in each admitted to ICU and with hospital length of stay (LOS) >1 day.

| Age | Sex | N | ICU | LOS >1d |
| --- | --- | --- | --- | --- |
| 18-35 | M | 105,205 | 1,004 (0.95%) | 2,212 (2.10%) |
|  | F | 131,676 | 1,014 (0.77%) | 2,255 (1.71%) |
| 35-45 | M | 58,986 | 1,506 (2.55%) | 3,228 (5.47%) |
|  | F | 73,396 | 1,265 (1.72%) | 3,072 (4.19%) |
| 45-55 | M | 70,767 | 3,060 (4.32%) | 6,543 (9.25%) |
|  | F | 86,373 | 2,283 (2.64%) | 5,620 (6.51%) |
| 55-60 | M | 36,319 | 2,056 (5.66%) | 4,349 (11.97%) |
|  | F | 41,947 | 1,517 (3.62%) | 3,692 (8.80%) |
| 60-65 | M | 31,125 | 2,104 (6.76%) | 4,244 (13.64%) |
|  | F | 34,445 | 1,585 (4.60%) | 3,677 (10.67%) |
| 65-70 | M | 10,477 | 844 (8.06%) | 1,800 (17.18%) |
|  | F | 9,447 | 578 (6.12%) | 1,433 (15.17%) |

The study consisted of two Boolean primary outcomes: 1) ICU admission; and 2) a hospital length of stay greater than 1 day. Across the entire study cohort, ICU admission rate was 2.67% (2.72%) and hospital length of stay greater than 1 day was 6.00% (6.10%). Both outcomes were strongly associated with age (p<2×10^−16^ each). Within each age strata, sex was also associated with both outcomes (all p<0.05), with men 41% (45%) more likely to be admitted to the ICU and 27% (29%) more likely to have a length of stay greater than 1 day than women, on average across age strata. Outcome rates across age and sex are included in Table 1.

### Chronic disease health history

Univariate associations were performed between both study outcomes and health history generated from health care claims from 26 common chronic diseases defined by the Chronic Conditions Data Warehouse across each age-sex strata. Analyses were omitted if the number of patients with the disease and with the study outcome did not exceed 20 within the strata. These sample sizes are included in Supplemental Table 1. Generally, for most chronic diseases that met sample size thresholds, there was a positive association between the presence of that condition and severe COVID-19 infection – particularly for patients early in life, but less pronounced for older patients (Figure 1). From these trends, we define four classes of chronic disease based on the observed effects and when they occur in life, though they certainly lie along a spectrum.

**Figure 1.**
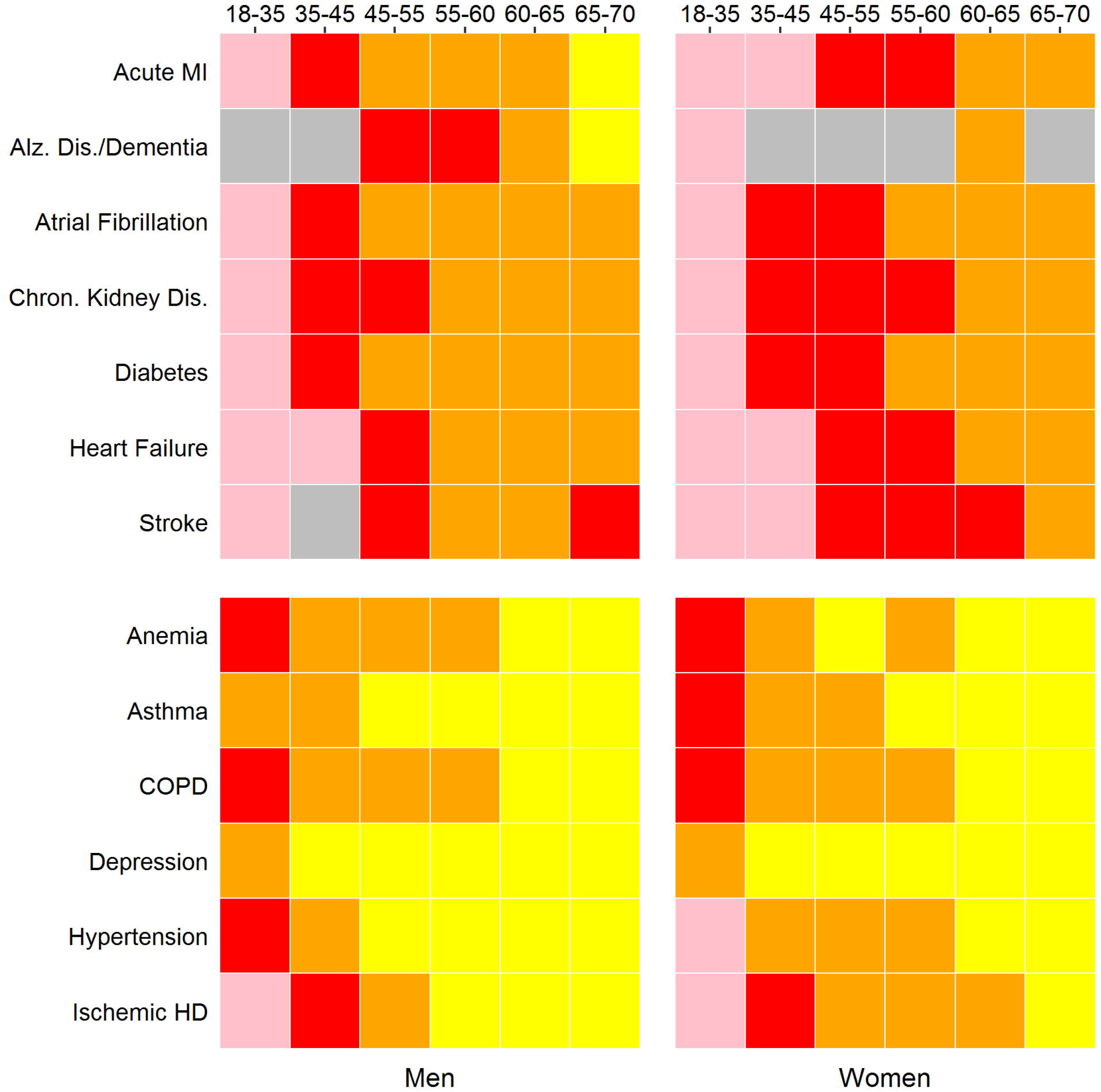
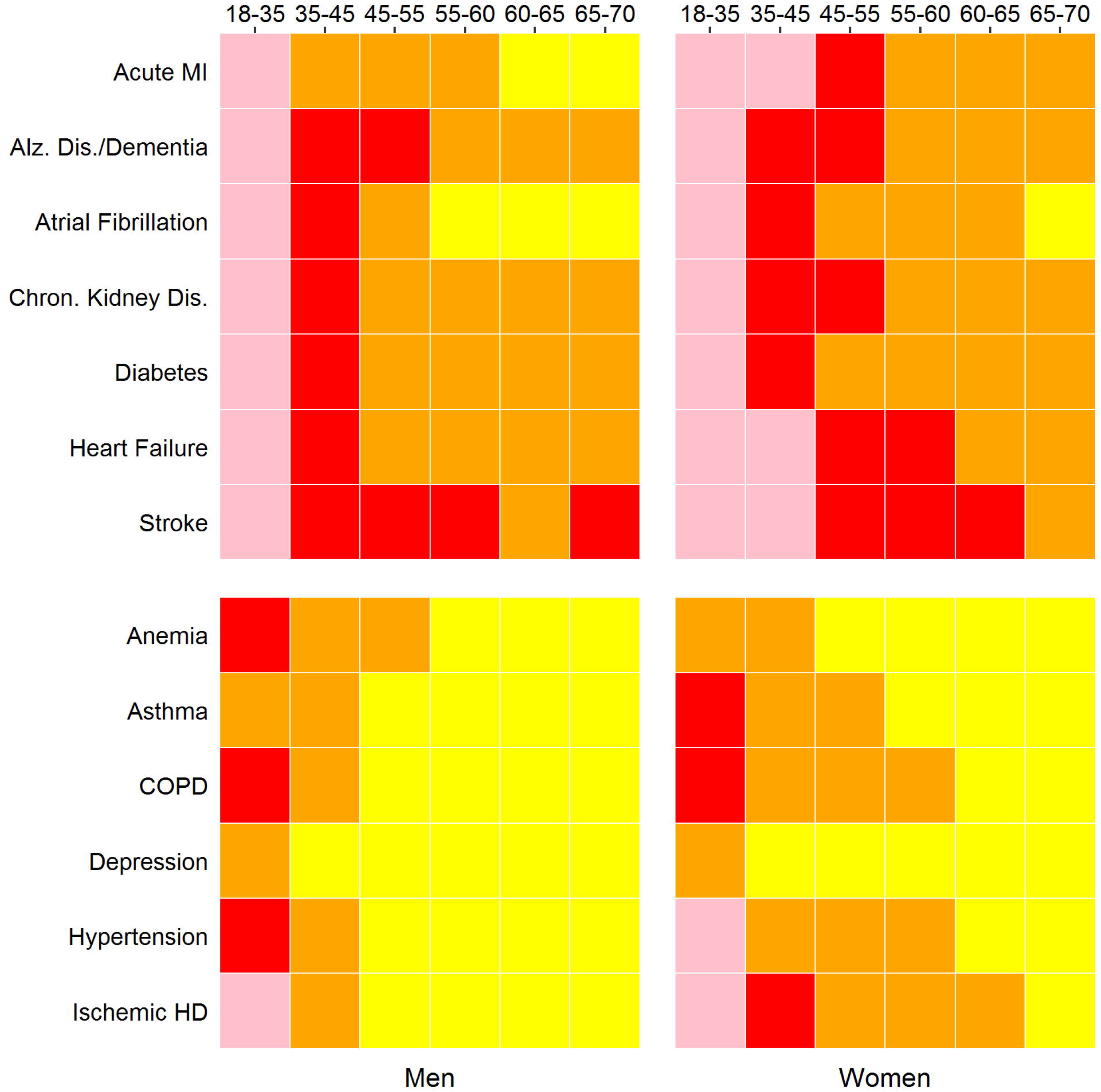
Effects of pre-existing health conditions on COVID-19 prognosis outcomes: A) ICU admission; B) hospital length of stay >1 day. Univariable odds ratio: pink: OR>8; red: OR>4; orange: OR>2; yellow: OR>1.2; gray: low sample size.

## Class 1

Class 1 diseases are defined by having substantial risk for both outcomes across age-sex strata. There are 7 diseases presented that meet this criteria: acute myocardial infarction, Alzheimer’s disease/senile dementia, atrial fibrillation, chronic kidney disease, diabetes, heart failure, and stroke. Among these, stroke is perhaps the most serious as the risk remains substantial (odds ratio >4) across most age-sex strata, between both COVID-19 prognosis outcomes. Among men and women ages 55-60, the odds of ICU admission among patients with a prior stroke is increased by a factor of 4.2 and hospital stay greater than 1 day by 4.4 (both p<2×10^−16^). Stroke does, however, remain a comparatively uncommon condition, particularly early in life. Chronic kidney disease and diabetes are, alternatively, very common with up to and exceeding 1000s of patients with both – these conditions and COVID-19 prognosis outcomes – in numerous strata, including earlier in life. Odds of ICU admission among all patients age 45-55 with chronic kidney disease is increased by a factor of 4.6, and 6.1 among patients 35-45. Similarly, the odds of ICU admission among diabetics in these age ranges are increased by a factor of 3.8 and 5.2, respectively (all p<2×10^−16^). Alzheimer’s disease and other forms of dementia have a sizable impact on COVID-19 prognosis outcomes, increasing the odds of ICU admission by a factor of 2.3 in patients 60-65 (p=8.1×10^−10^). Heart failure is, perhaps, second to stroke in terms of risk for severe infection while being 3-4 times more common. Patients 45-55 with heart failure have 5.8 increased odds of ICU admission and 4.5 of a hospital stay greater than 1 day (both p<2×10^−16^) while being more pronounced in women (p=1.7×10^−4^). The effect of acute myocardial infarction on COVID-19 prognosis is also more pronounced in women than men (p=5.4×10^−4^) with an increased odds of ICU admission among women 45-55 equal to 6.4 and 3.6 in men (both p<2×10^−16^). Lastly, among Class 1 disease, atrial fibrillation had the mildest average risk for severe COVID-19 infection, with an increase in the odds of ICU admission equal to 3.0 among sexes 55-60.

## Class 2

Class 2 diseases demonstrate similar trends to Class 1 diseases, but with milder risks in mid-and late-life. For these diseases, the increased odds of either severe COVID-19 outcome are typically near or below 2 across age-sex strata. These include anemia, asthma, chronic obstructive pulmonary disease, depression, hypertension, and ischemic heart disease. Notably, hypertension and ischemic heart disease have more pronounced effects in women than men, with hypertensive women 45-55 with having 2.7 increased odds of ICU admission compared to 2.0 in men (p=1.8×10^−8^) and same aged women with ischemic heart disease 3.4 compared to 2.2 in men (3.5×10^−6^). These diseases could be considered Class 1 in women.

## Class 3

Class 3 diseases have little to no impact on severe COVID-19 infection beyond early adult life. These include acquired hypothyroidism, cataract, glaucoma, hyperlipidemia, osteoporosis, rheumatoid and osteoarthritis, breast cancer in women, and prostate cancer and benign prostatic hyperplasia in men.

Too few observations were available to form meaningful conclusions from hip fracture, endometrial cancer, and lung cancer. Estimated risks for each disease across both outcomes within all age-sex strata are available in Supplemental Tables 2 & 3.

**Table 2.** AUC values from multivariable models for each age-sex strata and COVID-19 prognosis outcome.

| Age | Sex | ICU | LOS>1d |
| --- | --- | --- | --- |
| 18-35 | M | 0.799 (0.784, 0.814) | 0.786 (0.776, 0.796) |
|  | F | 0.821 (0.806, 0.836) | 0.808 (0.798, 0.818) |
| 35-45 | M | 0.720 (0.705, 0.734) | 0.694 (0.684, 0.705) |
|  | F | 0.749 (0.733, 0.764) | 0.735 (0.725, 0.745) |
| 45-55 | M | 0.700 (0.690, 0.711) | 0.668 (0.660, 0.675) |
|  | F | 0.735 (0.724, 0.746) | 0.707 (0.700, 0.715) |
| 55-60 | M | 0.705 (0.693, 0.717) | 0.663 (0.653, 0.672) |
|  | F | 0.711 (0.697, 0.725) | 0.692 (0.683, 0.701) |
| 60-65 | M | 0.685 (0.673, 0.697) | 0.667 (0.658, 0.676) |
|  | F | 0.706 (0.692, 0.719) | 0.687 (0.677, 0.696) |
| 65-70 | M | 0.687 (0.667, 0.706) | 0.665 (0.651, 0.679) |
|  | F | 0.712 (0.690, 0.734) | 0.689 (0.673, 0.704) |

## Multivariable analysis

Multivariable models were constructed using all disease indicators that met sample size thresholds within each age-sex strata, for each COVID-19 prognosis outcome. Parameter estimates for each strata are included in Supplemental Table 4. ROC curves were constructed from these models (Figure 2). There were a few general trends observed: 1) predictive determination generally decreased with increasing age; 2) within an age strata, prediction was always nominally greater in women compared to men; and 3) within an age-sex strata, prediction was always nominally stronger for ICU admission than length of stay >1 day. Thus, for a given age range, AUC values were largest for women when predicting ICU admission and lowest for men when predicting a length of stay >1 day. Mirroring the large disease effects observed in young adults, the models were most predictive among individuals 18-35, with AUC values ranging from 0.820 (women, ICU admission) to 0.784 (men, length of stay >1 day) in these cohorts. AUC values remain elevated 35-45 compared to older ages, at 0.746 and 0.721 for ICU admission for men and women, respectively. From 45-70 years, there appears to be few pronounced differences in prediction with AUC values hovering close to 0.7 for all other stratum/outcomes (Table 2).

**Figure 2.**
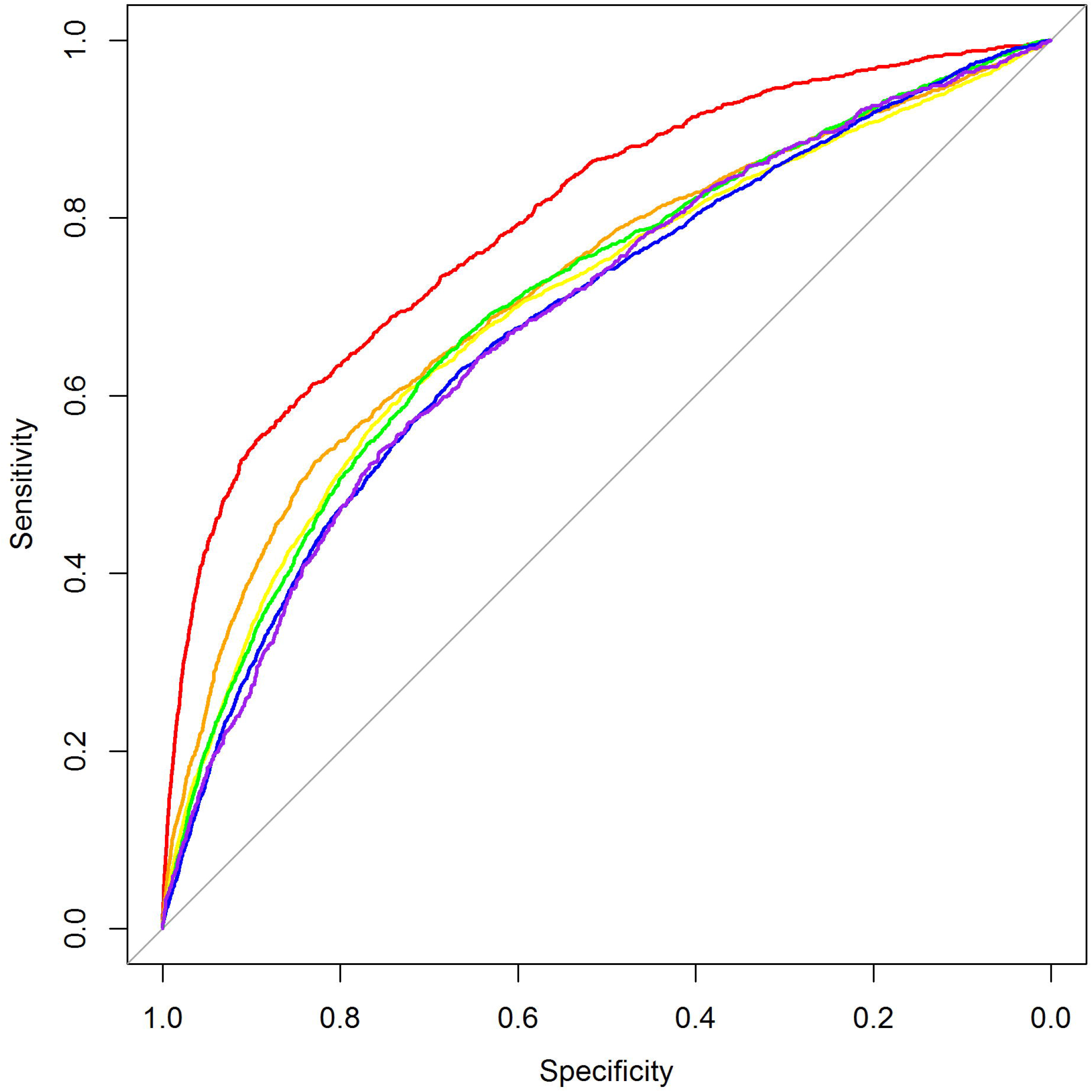
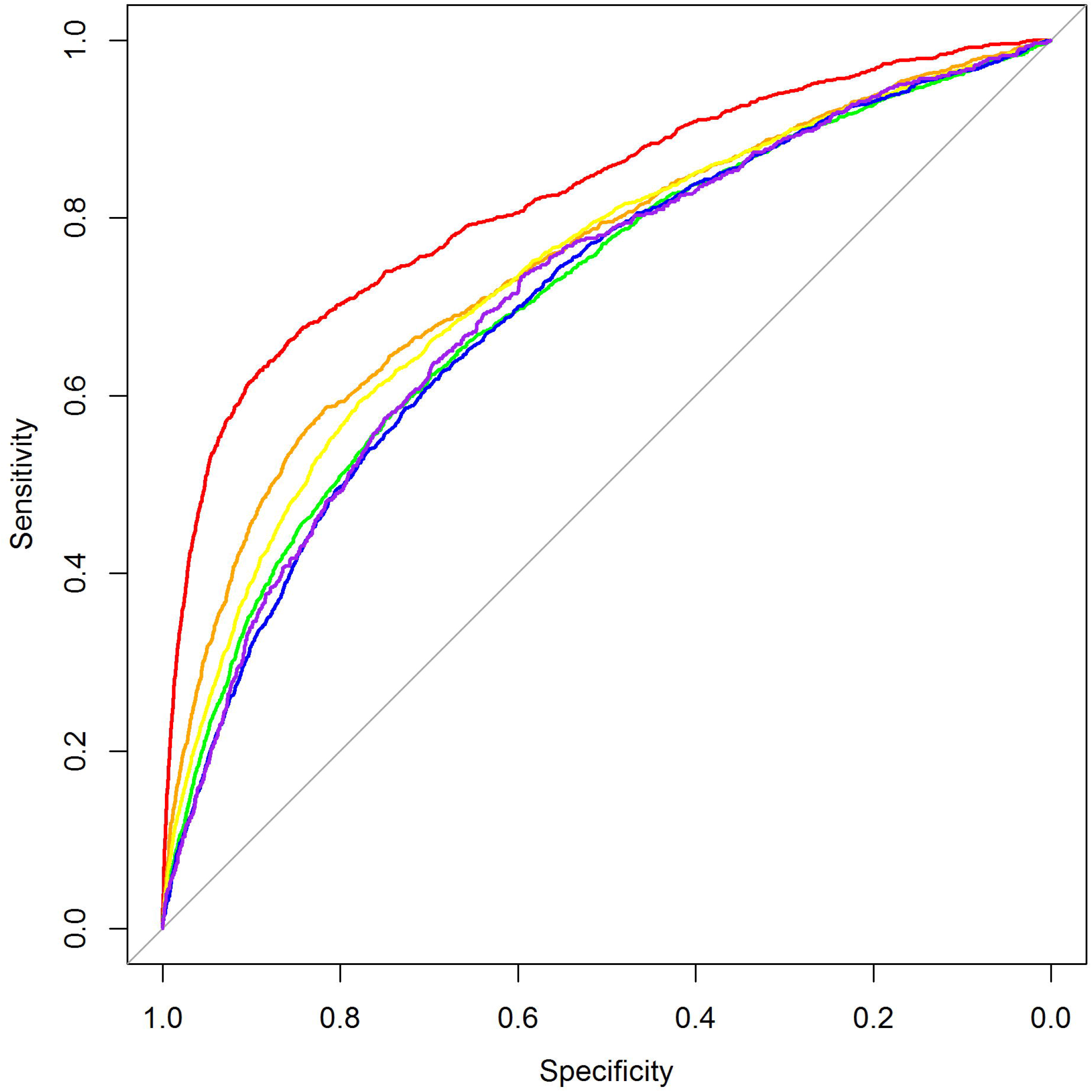
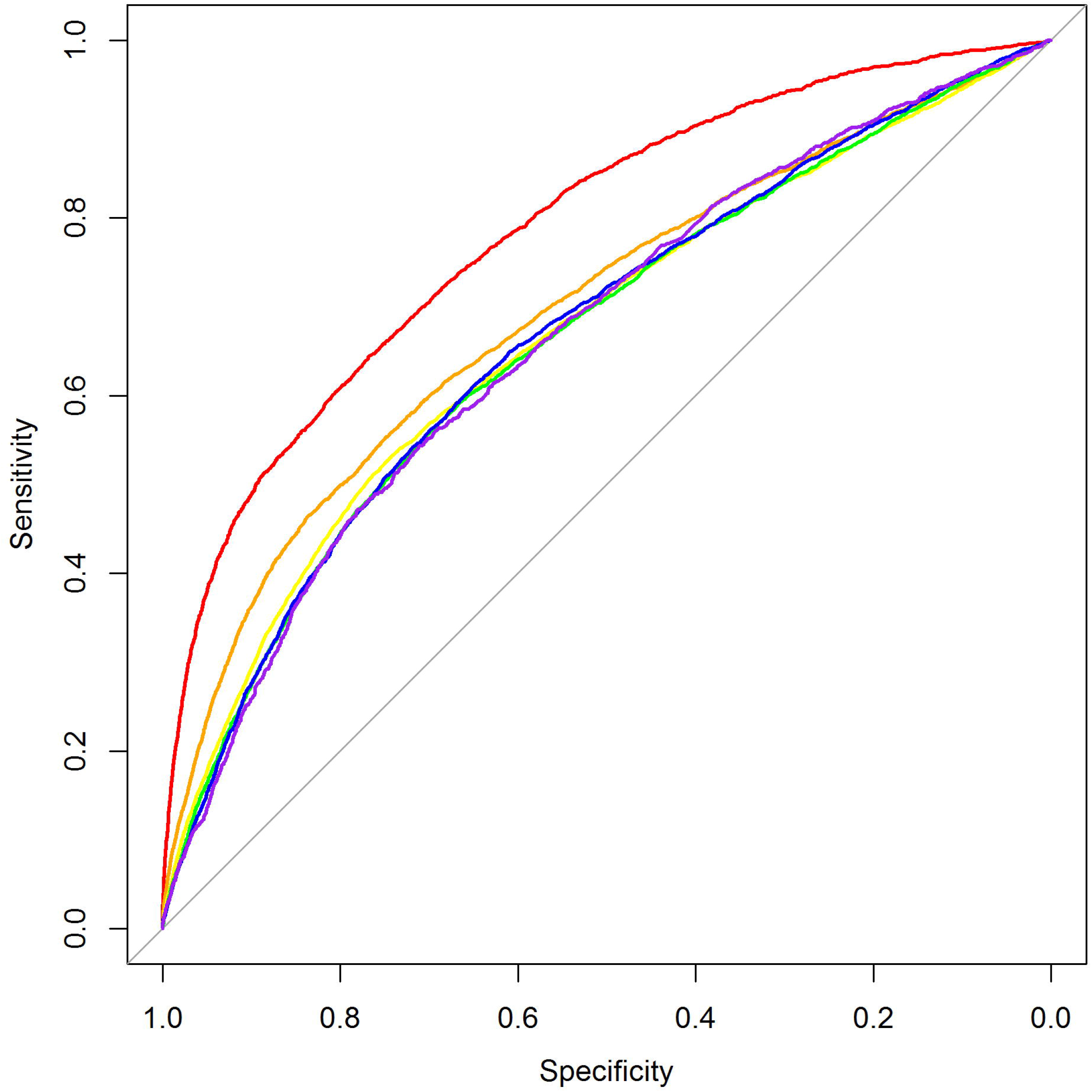
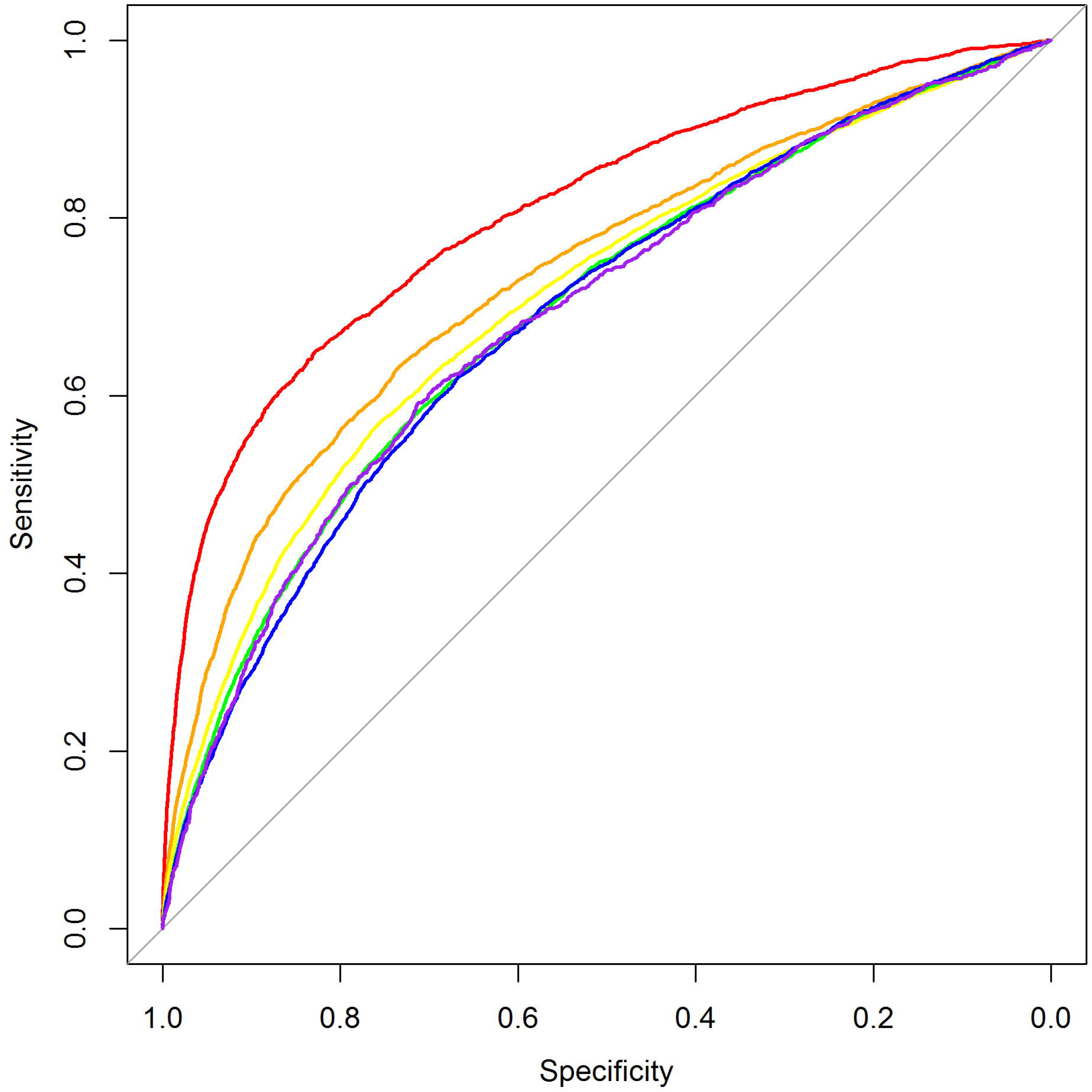
ROC curves from multivariable models. A) Men, ICU admission; B) Women, ICU admission; C) Men, hospital length of stay >1 day; D) Women, hospital length of stay >1 day. Red: 18-35; Orange: 35-45; Yellow: 45-55; Green: 55-60; Blue: 60-65; Purple: 65-70.

The likelihood an individual infected with COVID-19 has one of these severe outcomes can be estimated from the model parameters, and that probability can be calculated assuming the sample is representative of that individual (i.e., commercially insured, makes a health insurance claim with COVID-19). For example, the probability a 30-year old woman infected with COVID-19 with no pre-existing health conditions would have a 0.42% chance of being admitted to the ICU. Meanwhile, such a woman with asthma and diabetes would see that risk rise to 2.05%. An online risk calculator to estimate ICU admission rates based on pre-existing health conditions using this model is available at https://nwineinger.shinyapps.io/covid19_risk/.

Because pre-existing health conditions are generally not independent, multivariable analyses reveal conditional effects of disease which may better reveal the drivers of severe COVID-19 infection.

Among the Class 1 diseases, chronic kidney disease, diabetes, and stroke are conditionally associated with ICU admission across all age-sex strata. While the other Class 1 diseases generally demonstrate similar trends, there are some exceptions (negative coefficients): acute MI not associated in men under 45, Alzheimer’s disease/dementia in men 65-70, atrial fibrillation in women 35-45, heart failure in men 65-70. Among Class 2 diseases, positive coefficients are observed in all age-sex strata for asthma, chronic obstructive pulmonary disease, and hypertension. Anemia (women 65-70) and depression (women 55-60) both have negative coefficients in one age-sex strata, and ischemic heart disease, two (men 60-65, 65-70). Among interactions modeled between the four most common conditions (chronic kidney disease, diabetes, hyperlipidemia, and hypertension), negative coefficients were often observed (i.e., lessening of compounding disease effects). All model coefficients are included in Supplemental Table 4.

## Discussion

From 690,000 Americans, we defined classes of pre-existing health conditions with varying levels of severity risks across age-sex strata. In particular, we were able to quantify risks in young adults with low rates of severe infection and pre-existing conditions. Generally, the impact of disease in young adults was far greater, and was even large among diseases that showed little effect later in life – suggesting any pre-existing health condition in this age range is a strong indicator of susceptibility to the virus. The multivariable models we developed were most capable of predicting outcomes in 18-35 year olds, and should be strongly considered when distributing vaccines (or getting oneself to a vaccination site) within this age range. The freely accessible online risk calculator we developed from our models can be used to help guide these decisions. There users can calculate rate of ICU admission based on pre-existing health conditions and make comparisons with other risk factors or the population more broadly.

While various COVID-19 prognosis prediction models have been published,^10^ ours is the largest study solely from the U.S. population. This allows us to more accurately estimate risks for rare events (e.g., disease in young people). Furthermore, because Blue Cross Blue Shield is a network of companies that offer typically employer-purchased, commercial health insurance across the country, our data better represents the general population targeted in later stages of the vaccine rollouts. Our analysis strategy was to stratify the cohort based on age and sex, acknowledging these, particularly age, as well-established risk factors and recognizing rates of disease can differ drastically between groups. Previously published studies similar to ours have instead pooled their cohorts and used these as predictive features.^11,18,19^ Because age is such an overpowering risk factor, these models do not allow for much nuanced risk assessment in younger adults. For example, using another online risk calculator available,^18^ a healthy 38-year old male and another smoker with asthma, chronic obstructive pulmonary disease, and controlled diabetes would binned into identical risk tiers – one estimate provided even places the healthy individual at higher mortality risk. These models are not well calibrated for younger adults. Our model, instead, estimates the ICU admission rates of these example individuals at 1.3% and 8.2%, respectively – the latter being higher than the average risk of a 65 year-old man.

As vaccines become available to the general population but remain limited, important public health and personal decisions will need to be made. Who remains at high risk for a severe infection? Should age remain the main barometer? How does one compare the risks of a healthy 55-year old to a 35-year old with multiple health conditions? Should you get in line as soon as you are eligible, or would you wait and let others with more serious health concerns go first? While we encourage individuals to use the risk calculator we included to assess their personal risk, Figure 3 can provide general guidance on a population level. Here we present the median risk of ICU admission based on the number of Class 1 and Class 2 pre-existing conditions an individual may carry for each age-sex strata, up to age 65. While age remains perhaps the most relevant risk factor, there certainly are instances where younger individuals would be considered at higher risk. Vaccination strategies should consider these factors moving forward.

**Figure 3.**
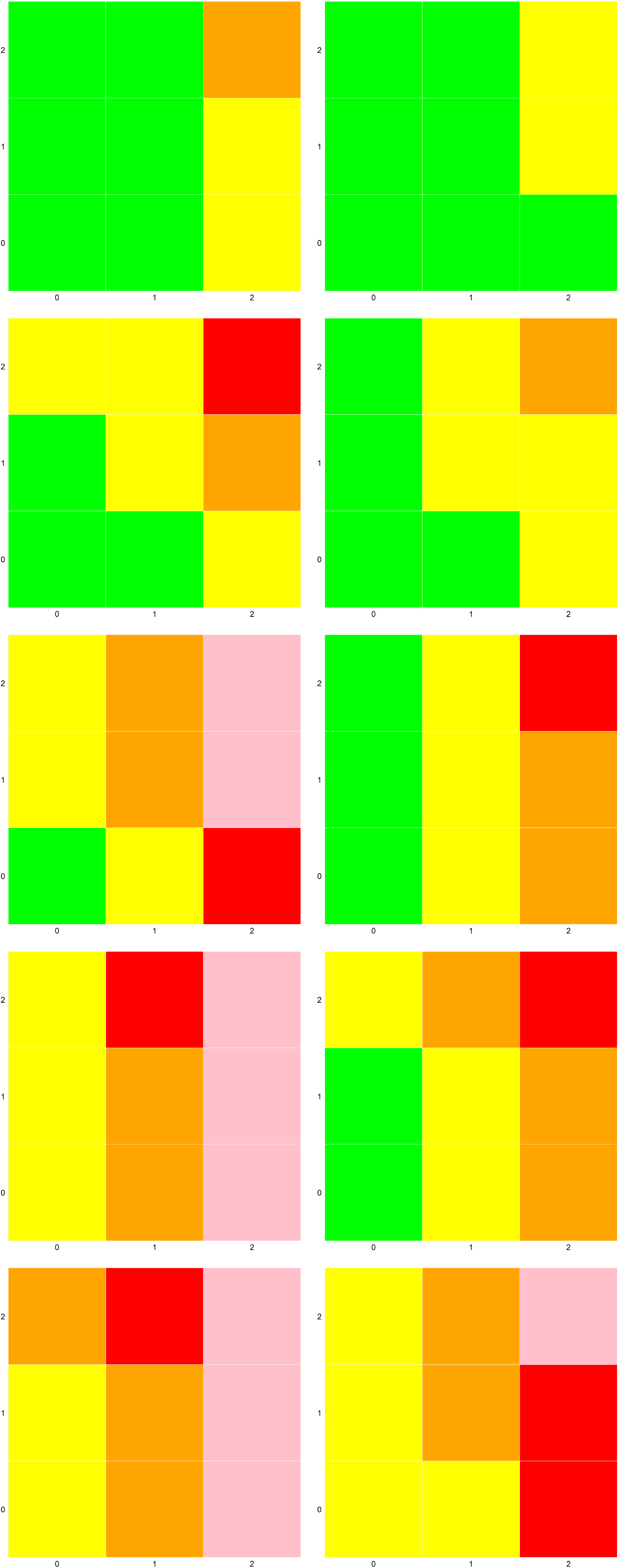
Median risk of ICU admission based on the number of Class 1 (x-axis) and Class 2 (y-axis) pre-existing conditions. Men (left), women (right). Top to bottom: ages 18-35, 35-45, 45-55, 55-60, 60-65. Colored according to probabilistic risks estimated from multivariable models: Green: 0-2.5%; Yellow: 2.5-5%; Orange: 5-7.5%; Red: 7.5-10%; Pink: >10%.

A strength of our study, in addition to the sheer size, is the commercially insured patients studied is more representative of the mostly healthy population that will be targeted in Phase 3 of the vaccination rollout. While such individuals are generally at low risk, the majority of the population – and thus the majority of vaccinations – falls in this category. Quantifying risks tiers among these individuals is critical while vaccine supply remains finite. However, it is not clear how the lack of data on individuals from the general population without commercial insurance would impact our results. Nor are individuals infected with COVID-19 that forego health care represented. Lastly, some other important subgroups, such as those immunocompromised, are considered high risk for severe infection, but were not directly included in our results.

Our hope is that individuals and public health policy makers can use the data presented in this study to make informed decisions on vaccinations for oneself and the general population. While vaccines remain in limited supply, the most vulnerable among us should receive priority. Our study provides insights into the risk factors among individuals at otherwise low risk.

## Data Availability

Data is the owned by Blue Cross Blue Shield Association and not available for distribution.

